# COVID-19 Infection Risk amongst 14,104 Vaccinated Care Home Residents: A national observational longitudinal cohort study in Wales, United Kingdom, December 2020 to March 2021

**DOI:** 10.1101/2021.03.19.21253940

**Authors:** Joe Hollinghurst, Laura North, Malorie Perry, Ashley Akbari, Mike B Gravenor, Ronan A Lyons, Richard Fry

**Affiliations:** Population Data Science and Health Data Research UK, Swansea University; Public Health Wales; Swansea University Medical School, Swansea University

## Abstract

**Background:** Vaccinations for COVID-19 have been prioritised for older people living in care homes. However, vaccination trials included limited numbers of older people.

**Aim:** We aimed to study infection rates of SARS-CoV-2 for older care home residents following vaccination and identify factors associated with increased risk of infection.

**Study Design and Setting:** We conducted an observational data-linkage study including 14,104 vaccinated older care home residents in Wales (UK) using anonymised electronic health records and administrative data.

**Methods:** We used Cox proportional hazards models to estimate hazard ratios (HRs) for the risk of testing positive for SARS-CoV-2 infection following vaccination, after landmark times of either 7 or 21-days post-vaccination. We adjusted hazard ratios for age, sex, frailty, prior SARS-CoV-2 infections and vaccination type.

**Results:** We observed a small proportion of care home residents with positive PCR tests following vaccination 1.05% (N=148), with 90% of infections occurring within 28-days. For the 7-day landmark analysis we found a reduced risk of SARS-CoV-2 infection for vaccinated individuals who had a previous infection; HR (95% confidence interval) 0.54 (0.30,0.95), and an increased HR for those receiving the Pfizer-BioNTECH vaccine compared to the Oxford-AstraZeneca; 3.83 (2.45,5.98). For the 21-day landmark analysis we observed high HRs for individuals with low and intermediate frailty compared to those without; 4.59 (1.23,17.12) and 4.85 (1.68,14.04) respectively.

**Conclusions:** Increased risk of infection after 21-days was associated with frailty. We found most infections occurred within 28-days of vaccination, suggesting extra precautions to reduce transmission risk should be taken in this time frame.

## Introduction

Vaccinations for SARS-CoV-2 in the UK have been prioritised to older people living in care homes [1], [2]. However, the efficacy of the COVID-19 vaccine in older people is relatively unknown, with very few trials recruiting older people and older people with frailty [3]. Specifically, the Oxford–AstraZeneca vaccine trials had less than 4% of participants over 70 years of age and those with comorbidities were a minority [4]. Similarly, the initial Pfizer BioNTECH trials included only 774 individuals aged 75 or over and 3,848 individuals aged 65 and over from a total of 18,198 vaccinated individuals [5].

Care homes are a keystone of adult social care. They provide accommodation and care for those needing substantial help with personal care, but more than that, they are people’s homes [6], [7]. In 2016, there were 11,300 care homes in the UK, with a total of 410,000 residents [8]. Within care homes people live in proximity, and may live with frailty and many different health conditions, making them susceptible to outbreaks of infectious disease [6]. COVID-19 is described by Lithande et al, as ‘…a dynamic, specific and real threat to the health and well-being of older people’ (2020,p.10) [9]. The impacts of COVID-19 on this sub-population have been reported widely in both international and UK media, and in a growing peer reviewed literature.

Here, we produced a rapid report, in near real-time, on the risk of SARS-CoV-2 infection for vaccinated care home residents. This is the first study we are aware of investigating this vulnerable sub-population. Furthermore, we included information on previous positive SARS-CoV-2 PCR tests, age, sex, and frailty. We were able to do this using the existing infrastructure and linked data from the Secure Anonymised Information Linkage (SAIL) Databank [10]–[12].

## Methods

### Study design and setting

We conducted an observational data-linkage study for older care home residents in Wales (UK). We used data on 14,104 individuals receiving a SARS-CoV-2 vaccination from 4^th^ December 2020 to 12^th^ February 2021 and testing data from 4^th^ December 2020 to 4^th^ March 2021 to investigate positive SARS-CoV-2 PCR tests following a vaccination.

### Data sources

We used linked longitudinal data from the SAIL Databank to create our datasets [10]–[12]. Specifically, we used the COVID Vaccine Dataset (CVD) to identify individuals living in care homes who had received a vaccination. We included all individuals identified as an ‘older adult resident in a care home’. The Pathology COVID-19 Daily (PATD) data was used to identify dates of positive SARS-CoV-2 PCR tests. A cleaned and pre-linked version of the Welsh Demographic Service Dataset (WDSD) was used to determine demographic information for each individual [13]. We also linked to the Patient Episode Database for Wales (PEDW) to include an indication of frailty.

### Hospital Frailty Risk Score

The Hospital Frailty Risk Score (HFRS) was developed using Hospital Episode Statistics (HES), a database containing details of all admissions, Emergency Department attendances and outpatient appointments at NHS hospitals in England, and validated on over one million older people using hospitals in 2014/15 [14]. The HFRS uses the International Classification of Disease version 10 [15] (ICD-10) codes to search for specific conditions from secondary care. A weight is then applied to the conditions and a cumulative sum is used to determine a frailty status of: Low, Intermediate or High. We additionally included a HFRS score of ‘No score’ for people who had not been admitted to hospital in the look back period. We calculated the HFRS using the Patient Episode Database for Wales (PEDW), the Welsh counterpart to HES, on the vaccination date, with a two year look back of all hospital admissions recorded in Wales.

### Variables

Our outcome of interest was the time to a positive SARS-CoV-2 PCR test following a vaccination. Individuals were censored for death or the end of study period. We included covariates for previous positive SARS-CoV-2 PCR tests (yes/no), age (continuous), sex (male/female), frailty (HFRS: no-score, low, intermediate, high), and vaccine manufacturer (Oxford-AstraZeneca, Pfizer-BioNTECH). Previous positive SARS-CoV-2 PCR tests were identified at any time point before vaccination.

### Statistical methods

We included basic demographic information and investigated differences between individuals who had a positive SARS-CoV-2 PCR test following vaccination. We produced a Kaplan-Meier survival curve and an empirical cumulative distribution function for the time to first positive SARS-CoV-2 PCR test following vaccination. We also calculated Hazard Ratios (HR) for our covariates using univariable and multivariable Cox proportional hazards models. For the Cox proportional hazards models we defined two landmark periods for immunisation, 7 and 21-days. In a landmark analysis, only those who have not had an event (positive PCR test) for the specified time period are included. In other words, individuals who had a positive SARS-CoV-2 PCR test within the 7 and 21-day periods were removed from the respective analyses, see Figure S1 for an example. We included these periods as a proxy for the varying number of days for the vaccine to become effective. As a sensitivity analysis we repeated the analysis with the 7 and 21-day immunisation (landmark) periods, but applied a maximum follow-up of 14-days. Individuals were right censored for death or the end of the study period, whichever occurred first. Violations of the proportional hazards assumption were tested for using Schoenfeld residuals.

## Results

We identified 14,501 vaccinated older adult residents in a care home in the CVD dataset. We removed 240 residents prior to analysis due to incomplete demographic information. We restricted the age group to those aged 60+, removing a further 157 individuals, resulting in 14,104 residents used for analysis. The basic demographic information for the total cohort and the cohort stratified by those who had a subsequent positive SARS-CoV-2 PCR test following vaccine is presented in Table 1. Figure 1 shows the Kaplan-Meier curve for the time to first positive PCR test following vaccination. The curve indicates an overall small proportion of individuals testing positive following vaccination. Figure 2 shows the empirical cumulative distribution function for the times between first positive PCR test and vaccination. The Kaplan-Meier curve and empirical cumulative distribution function suggest a susceptible period of vaccinated individuals up to 42 days, with approximately 40% of individuals having a positive PCR test within 7 days, 60% within 14-days, 85% within 21-days, 90% within 28-days, and over 95% within 35-days.

**Table 1.**
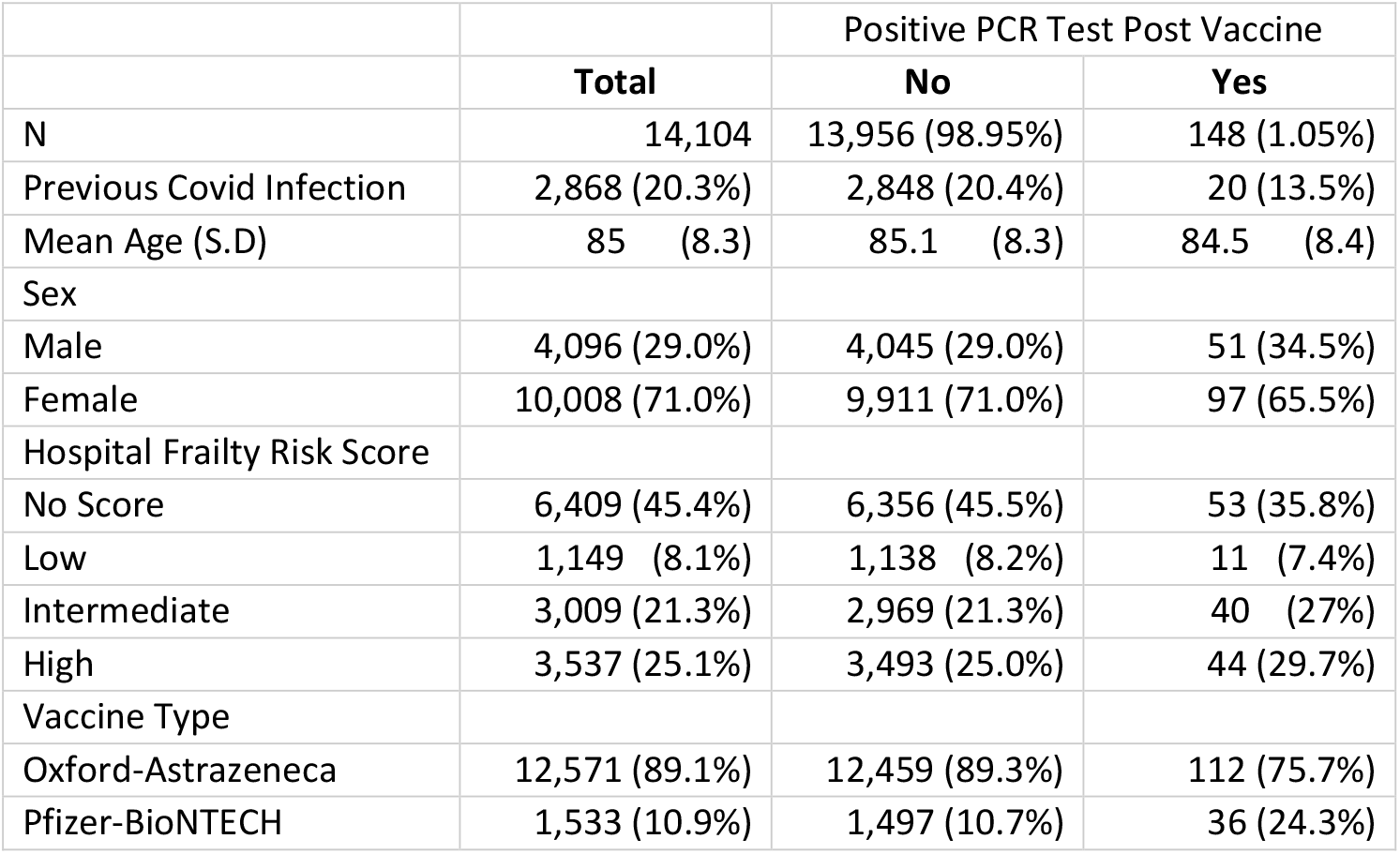
Demographic information for the total cohort and the cohort stratified by those who had a subsequent positive SARS-CoV-2 PCR test following vaccination.

**Figure 1.**
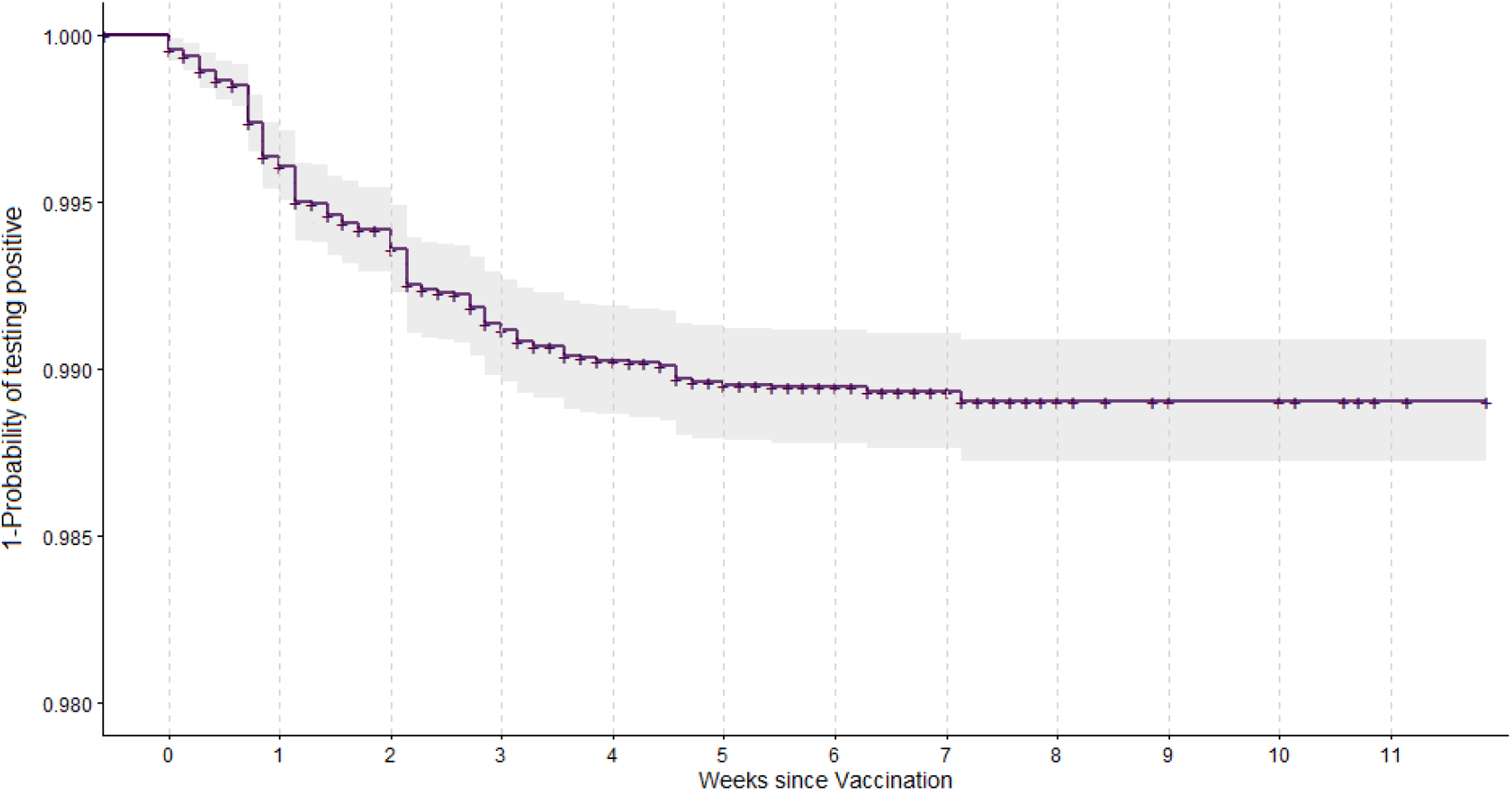
Kaplan-Meier curve for the time to the first positive SARS-COV-2 PCR test following vaccination.

**Figure 2.**
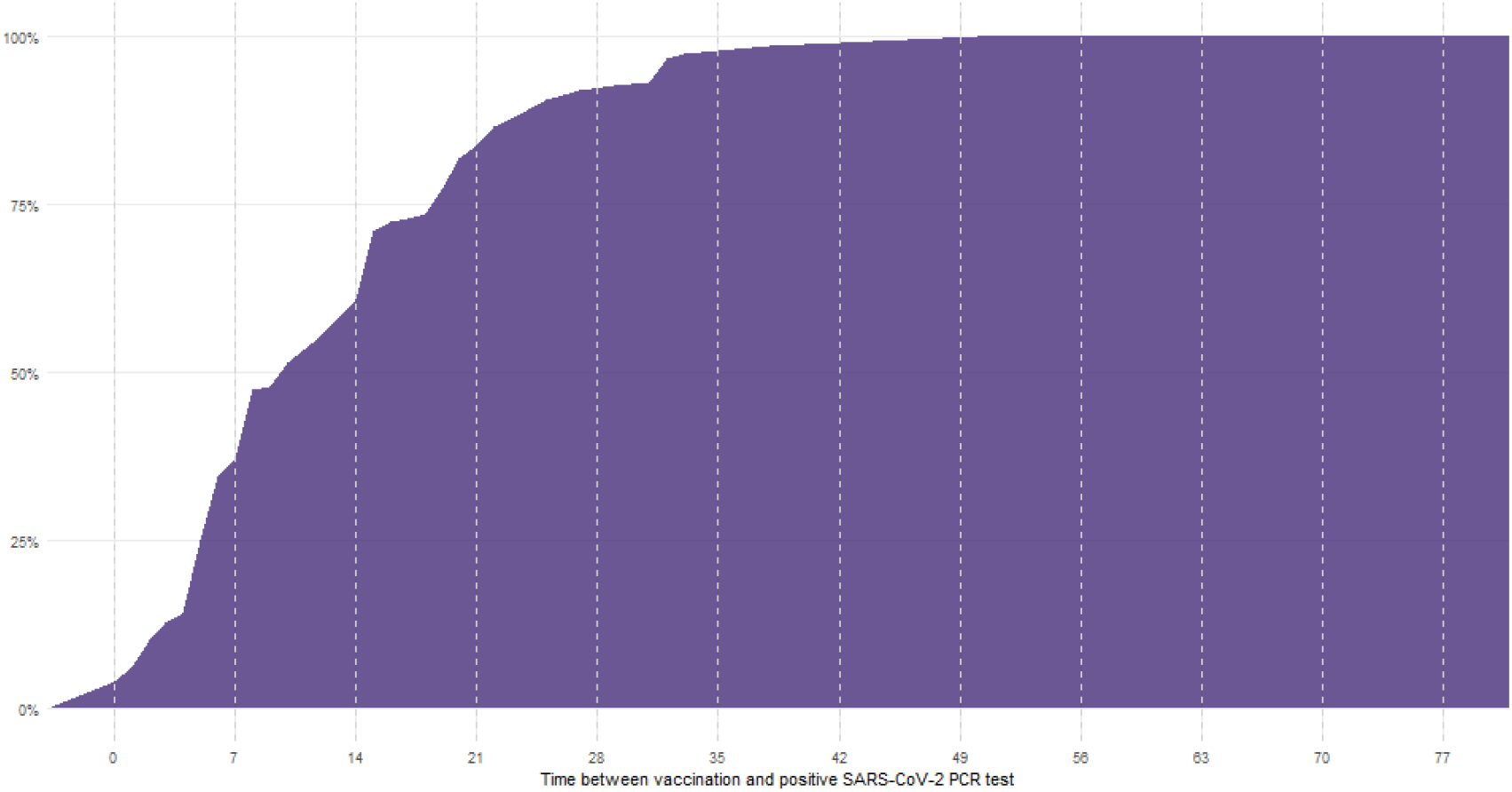
Cumulative Distribution Function (CDF) for the time between vaccination and first positive PCR test (N = 148).

Hazard Ratios and 95% confidence intervals for the Cox proportional hazards models are presented in Table 2 and Table 3. In our multivariable analyses, we found a reduced risk of SARS-CoV-2 infection for vaccinated individuals who had a previous infection after a 7-day immunisation period; HR (95% confidence interval) 0.54 (0.30,0.95), and an increased risk for those receiving the Pfizer-BioNTECH vaccine HR 3.83 (2.45,5.98). The 21-day immunisation period multivariable model indicated frailty as a risk factor, with low frailty having a HR of 4.59 (1.23,17.12) and intermediate frailty with a HR of 4.85 (1.68,14.04). The Schoenfeld residual test indicated only the 21-day immunisation / 14-day observation model deviated from the proportional hazards assumption at the 95% level (p-value 0.04), all other models met proportional hazards assumptions.

**Table 2.** Univariable and multivariable Cox Proportional Hazards models with 7 and 21-day landmark times applied to the cohort. Hazard ratios and presented with 95% confidence intervals. Results that are statistically significant at the 95% level are highlighted in bold font.

|  | 7-day landmark time |  | 21-day landmark time |  |
| --- | --- | --- | --- | --- |
| HRs (95% CI) | Univariable | Multivariable | Univariable | Multivariable |
| Age | 1.004 (0.98,1.028) | 1.01 (0.985,1.036) | 1.024 (0.976,1.074) | 1.031 (0.981,1.084) |
| Previous positive PCR test (Reference: No) |  |  |  |  |
| Yes | 0.667 (0.379,1.175) | **0.535 (0.301,0.949)** | 0.931 (0.353,2.46) | 0.807 (0.301,2.159) |
| Sex (Reference: Female) |  |  |  |  |
| Male | 1.39 (0.919,2.104) | 1.428 (0.929,2.195) | 1.246 (0.56,2.773) | 1.285 (0.563,2.932) |
| Hospital Frailty Risk Score (Reference: No score) |  |  |  |  |
| Low | 1.597 (0.791,3.226) | 1.605 (0.794,3.244) | **4.487 (1.205,16.71)** | **4.585 (1.229,17.12)** |
| Intermediate | 1.645 (0.996,2.717) | **1.767 (1.066,2.927)** | **4.662 (1.62,13.417)** | **4.852 (1.677,14.04)** |
| High | 1.298 (0.777,2.168) | 1.374 (0.82,2.303) | 2.511 (0.797,7.913) | 2.574 (0.813,8.15) |
| Vaccine (Reference: Oxford Astrazeneca) |  |  |  |  |
| Pfizer-BioNTECH | **3.362 (2.166,5.216)** | **3.829 (2.452,5.979)** | 1.921 (0.727,5.077) | 2.196 (0.821,5.873) |
| - | - | - | - | - |
| Observations | 13,989 | 13,989 | 13,605 | 13,605 |
| Events | 97 | 97 | 27 | 27 |
| - | - | - | - | - |
| Concordance |  | 0.669 (s.e. 0.028 ) |  | 0.693 (s.e. 0.052 ) |
| Global Schoenfeld residual |  |  |  |  |
| Chi-squared |  | 8.9 |  | 7.8 |
| Degrees of freedom |  | 7 |  | 7 |
| p-value |  | 0.26 |  | 0.35 |

**Table 3.**
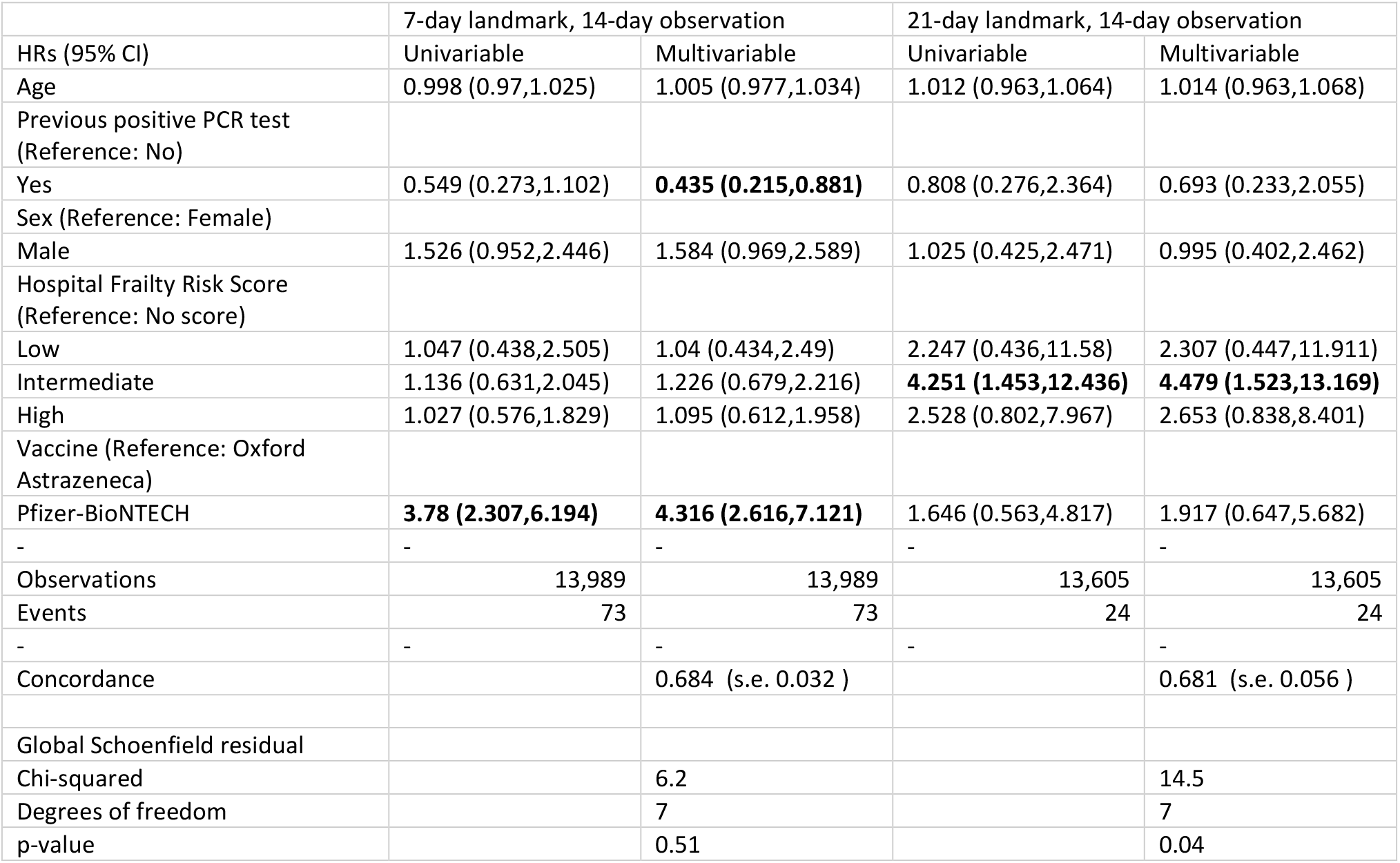
Univariable and multivariable Cox Proportional Hazards models with 7 and 21-day landmark times applied to the cohort and a maximum 14-day observation period. Hazard ratios and presented with 95% confidence intervals. Results that are statistically significant at the 95% level are highlighted in bold font.

## Discussion

Our study focussed on older adults resident in care homes. This is a particularly vulnerable sub-population that has not previously been studied in relation to infection rates for SARS-CoV-2 following vaccination, and which has suffered considerably from the most severe effects of the pandemic. Our study used a large cohort of 14,104 individuals, which is comparable to the entire case population of the ChAdOx1 nCoV-19 vaccine (N = 12,174) [4]. We were also able to include information on frailty, previous infections and vaccination received.

We found 148 (1.05%) of individuals in our cohort had a positive PCR-test following vaccination. The Kaplan-Meier curve and empirical cumulative distribution function suggest a susceptible period for infection of up to 42-days, with approximately 99% of infections occurring within this period. It is well known that there is a delay following immunisation for the vaccine to be effective, but this highlights the need for extended vigilance during this period for this highly vulnerable care home population.

We found a large, and statistically significant reduced risk for infection post vaccine for individuals who had already had a SARS-CoV-2 infection. The risk was approximately halved, and those who had a prior infection may be more robust and have existing antibodies, leading to a reduced risk to subsequent infections. Increased levels of frailty, determined by the HFRS, were associated with substantially increased risk of infection post vaccine (up to almost 5-fold increase for intermediate frailty in the 21-day landmark post vaccination). Frailty is complex, and those living with high levels of frailty may need additional support. In particular, increased care requires additional contact with carers, and subsequently an increased risk of transmission of SARS-CoV-2 compared to those who are more independent or can be isolated. We note that the effect was not quite as large in the highest frailty group. This simply may reflect uncertainty in the risk estimates, or may be the result of more complex management or identification of risk in this group.

In the 7-day landmark analysis there was evidence to suggest an increased risk of infection post vaccination for those receiving the Pfizer-BioNTECH vaccine compared to the Oxford-AstraZeneca vaccine, but no statistically significant difference in the 21-day landmark analysis. This suggests the Oxford-AstraZeneca vaccine has a potentially shorter time to become effective in our cohort.

### Strengths

This analysis was possible using the existing infrastructure of the SAIL Databank and we will continue to investigate adverse events for individuals receiving a vaccination. We were able to rapidly develop and analyse a large cohort of care home residents with the inclusion of individual level information.

### Limitations

Due to the nature of the vaccination rollout, we only had a limited follow-up time and subsequently a small number of events. At the time of analysis we did not include information on second doses due to very small numbers. We will continue to monitor the care home population and will update our analysis with an extended follow-up time and second doses when possible. In further work we aim to include background prevalence of COVID-19, and multi-morbidities and we will investigate additional adverse events such as mortality and hospitalisation.

### Conclusion

Our findings suggest care home residents with frailty are the most susceptible to infection post vaccination and should be prioritised for a second dose of the SARS-CoV-2 vaccine. We also found a susceptible period of reinfection of up to 42-days, indicating extra care and precautions should be taken in this period.

## Data Availability

The data used in this study are available in the SAIL Databank at Swansea University, Swansea, UK. All proposals to use SAIL data are subject to review by an independent Information Governance Review Panel (IGRP). Before any data can be accessed, approval must be given by the IGRP. The IGRP gives careful consideration to each project to ensure proper and appropriate use of SAIL data. When access has been approved, it is gained through a privacy-protecting safe haven and remote access system referred to as the SAIL Gateway. SAIL has established an application process to be followed by anyone who would like to access data via SAIL https://www.saildatabank.com/application-process.

## Acknowledgements

This work uses data provided by patients and collected by the NHS as part of their care and support. We would also like to acknowledge all data providers who make anonymised data available for research.

We wish to acknowledge the collaborative partnership that enabled acquisition and access to the de-identified data, which led to this output. The collaboration was led by the Swansea University Health Data Research UK team under the direction of the Welsh Government Technical Advisory Cell (TAC) and includes the following groups and organisations: the Secure Anonymised Information Linkage (SAIL) Databank, Administrative Data Research (ADR) Wales, NHS Wales Informatics Service (NWIS), Public Health Wales, NHS Shared Services and the Welsh Ambulance Service Trust (WAST). All research conducted has been completed under the permission and approval of the SAIL independent Information Governance Review Panel (IGRP) project number 0911.

We used the STROBE checklist to create this manuscript, an Explanation and Elaboration article discusses each checklist item and gives methodological background and published examples of transparent reporting. The STROBE checklist is best used in conjunction with this article (freely available on the Web sites of PLoS Medicine at http://www.plosmedicine.org/, Annals of Internal Medicine at http://www.annals.org/, and Epidemiology at http://www.epidem.com/). Information on the STROBE Initiative is available at www.strobe-statement.org.

## SUPPLEMENTARY MATERIAL

**Figure S1.**
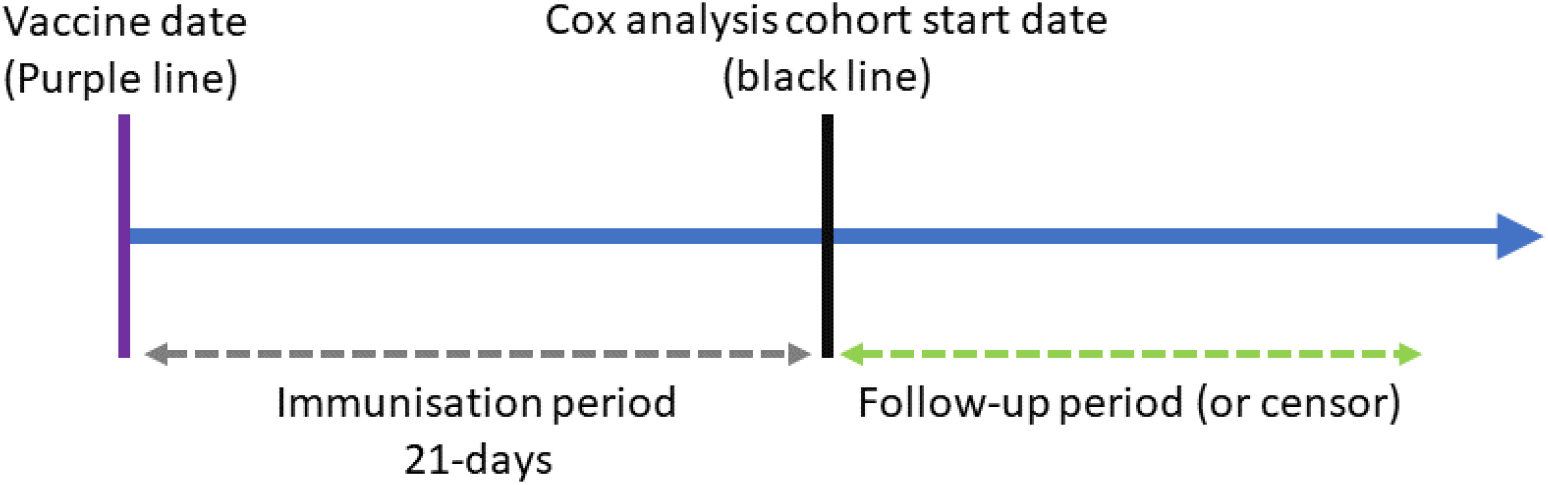
Immunisation (landmark) analysis timeline example.

## REFERENCES

[1] Public Health Wales, “Vaccination Priority List - Wales,” 2020. [Online]. Available: https://phw.nhs.wales/topics/immunisation-and-vaccines/covid-19-vaccination-information/eligibility-for-the-vaccine/. [Accessed: 01-Feb-2021].

[2] Public Health England, “Vaccination Priority List - England,” 2020. [Online]. Available: https://www.gov.uk/government/publications/covid-19-vaccination-care-home-and-healthcare-settings-posters/covid-19-vaccination-first-phase-priority-groups. [Accessed: 01-Feb-2021].

[3] R. L. Soiza, C. Scicluna, and E. C. Thomson, “Efficacy and safety of COVID-19 vaccines in older people,” Age Ageing, Dec. 2020.

[4] M. D. Knoll and C. Wonodi, “Oxford–AstraZeneca COVID-19 vaccine efficacy,” Lancet, vol. 397, no. 10269, pp. 72–74, Jan. 2021.

[5] F. P. Polack et al., “Safety and Efficacy of the BNT162b2 mRNA Covid-19 Vaccine,” N. Engl. J. Med., vol. 383, no. 27, pp. 2603–2615, Dec. 2020.

[6] D. Challis et al., “Dependency in older people recently admitted to care homes.,” Age Ageing, vol. 29, no. 3, pp. 255–260, 2000.

[7] A. L. Gordon et al., “Commentary: COVID in Care Homes—Challenges and Dilemmas in Healthcare Delivery,” Age Ageing, 2020.

[8] “Competitions and Markets Authority. (2017). Care homes market study. Final report. Competitions and Markets Authority, UK.”

[9] F. E. Lithander et al., “COVID-19 in Older People: A Rapid Clinical Review,” Age Ageing, 2020.

[10] D. V Ford et al., “The SAIL Databank: building a national architecture for e-health research and evaluation,” BMC Health Serv. Res., vol. 9, no. 1, p. 157, 2009.

[11] R. A. Lyons et al., “The SAIL databank: linking multiple health and social care datasets,” BMC Med. Inform. Decis. Mak., vol. 9, no. 1, p. 3, 2009.

[12] K. H. Jones et al., “A case study of the secure anonymous information linkage (SAIL) gateway: A privacy-protecting remote access system for health-related research and evaluation,” J. Biomed. Inform., 2014.

[13] J. Lyons et al., “Understanding and responding to COVID-19 in Wales: protocol for a privacy-protecting data platform for enhanced epidemiology and evaluation of interventions,” BMJ Open, vol. 10, no. 10, 2020.

[14] T. Gilbert et al., “Development and validation of a Hospital Frailty Risk Score focusing on older people in acute care settings using electronic hospital records: an observational study,” Lancet, 2018.

[15] W. H. Organization and others, “ICD-10: international statistical classification of diseases and related health problems: tenth revision,” 2004.

